# Challenges Hindering the Translation of Plant Anticonvulsants from Bench to Bedside: A Scoping Review

**DOI:** 10.1101/2024.11.13.24317300

**Authors:** Joana Opoku, Patrick Amoateng, Kennedy Kwami Edem Kukuia, Samuel Ankamah, Emelia Oppong Bekoe, Samuel Adjei, Dorcas Osei-Safo, Samuel Binamin Kombian

## Abstract

**Aim:** To identify the challenges that hinder the successful translation of anticonvulsants from plant origins, from preclinical research to clinical application.

**Design:** This review was conducted using the Joanna Briggs Institute (JBI) guidance for a scoping review.

**Data Sources:** The following bibliographic databases were searched between November 11th and 20th, 2023: PubMed, Scopus, Google Scholar and ClinicalTrials.gov. The search in Google Scholar was done via a third-party application called Harzing’s Publish or Perish, where the search results limit was set at 1000 relevant articles. After full-text review, the reference lists of the included articles were examined to identify additional sources.

**Review Methods:** The Population, Intervention, Comparison, Outcome, Time, Setting (PICOTS) framework was employed in developing the eligibility criteria. The articles were uploaded to Rayyan.ai for title and abstract screening, as well as full-text article review. Data was extracted and synthesised from included studies using a table.

**Results:** Thirty-seven articles met the eligibility criteria. From these articles, six distinct categories of challenges were identified: (1) methodologic issues; (2) insufficient evidence to support the use of herbal drugs in clinical settings; (3) financial disincentives; (4) challenges in obtaining natural products or its active principles; (5) poor pharmacokinetics; clinical trials and regulatory challenges.

**Conclusion:** Understanding and effectively addressing these challenges will ensure that more plant-based anticonvulsants are successfully translated into clinical practice, thereby enhancing the treatment of epilepsy. This review also made recommendations to tackle some of these challenges that plant anticonvulsants face in the drug development process.

## 1. INTRODUCTION

Epilepsy arises from increased neural excitability and an imbalance between excitatory and inhibitory signals, resulting in seizures. (Anwar et al., 2020). The International League Against Epilepsy (ILAE) defines epilepsy as a brain disorder characterised by any of the following: (1) two or more unprovoked (or reflex) seizures occurring more than 24 hours apart; (2) one unprovoked (or reflex) seizure with a high likelihood of further seizures (at least 60%) within the next 10 years, similar to the risk after two unprovoked seizures; or (3) the diagnosis of an epilepsy syndrome. Epilepsy is considered resolved in individuals who either had an age-dependent epilepsy syndrome and have surpassed the relevant age, or who have been seizure-free for the past 10 years, with at least 5 years without anti-seizure medication (Fisher et al., 2014). The World Health Organization estimates that approximately 50 million people around the world have epilepsy, positioning it as one of the most prevalent neurological conditions. Additionally, only up to 70% of individuals with epilepsy could achieve seizure control with proper use of anti-seizure medications (World Health Organization (WHO), 2019). This emphasises the need for effective anticonvulsants to help more patients attain a seizure-free status.

Although various antiseizure drugs (ASDs), whether used individually or in combination, are employed for the symptomatic treatment of epileptic seizures, approximately one-third of epilepsy patients experience seizures that are resistant to pharmacotherapy. Individuals with drug-resistant epilepsy (DRE) face heightened risks of premature death, injuries, psychosocial challenges, and a diminished quality of life, making the need for more effective treatments a critical clinical priority (Löscher et al., 2020). As such, many researchers are dedicated to discovering novel and effective treatments for epilepsy, and this includes exploring plant sources with anticonvulsant activities (Zhu et al., 2014).

Since ancient times, a variety of plants and their products have been used in the treatment of epilepsy in folkloric medicine. Their rich source of biological and chemical diversity cannot be denied (Quintans et al., 2008). Many reports have documented the efficacy of plants with anticonvulsant activity. For example, among the nearly 60 plants that are commonly used to treat epilepsy by Tanzanian traditional healers in Africa, *Abrus precatorius* L., *Clausena anisata* (Willd.) Oliv., and *Hoslundia opposita* Vahl. have shown definitive anticonvulsant activity from experimental literature reports (Moshi et al., 2005). Furthermore, among 15 herbal drugs used in traditional Chinese medicine, *Acorus tatarinowii* Schott., *Bupleurum chinense* DC., *Ligusticum chuanxiong* Hort., *Paeonia suffruticosa* Andr., and *Salvia miltiorrhiza* Bge. have demonstrated significant antiepileptic activity (Cao & Fang, 2006). Even though numerous plants have been studied, and proven to be effective in the treatment of epilepsy, most of them have not been successfully developed into standard medications (Manchishi, 2018). This necessitates the identification of the various reasons for the lack of successful assimilation or translation of most plant anticonvulsants from bench to bedside.

While the efficacy of many plant-derived anticonvulsants have been documented in various preclinical studies (Cao & Fang, 2006; Moshi et al., 2005), there is a significant gap in the literature addressing the specific challenges and barriers that hinder their translation from bench to bedside. Therefore, this study sought to identify the various challenges that hinder the successful translation of plant anticonvulsants from the laboratory setting to the clinical setting.

### 1.1 OBJECTIVES

The primary aim of this research was to identify the documented challenges that hinder the successful translation of natural plant anticonvulsants from preclinical research to clinical application. The research question being addressed was: “What are the key challenges preventing the effective transition of plant anticonvulsants, despite extensive research efforts, from the laboratory setting to clinical practice?”

## 2. METHODS

### 2.1 Protocol and Registration

The protocol for this study was drafted *a priori* (Opoku et al., 2024) based on the updated methodological guidance for the conduct of scoping reviews by JBI (Peters et al., 2020), and was revised by the authors. This review followed the guidelines of the PRISMA extension for Scoping Reviews (PRISMA-ScR) (Tricco et al., 2018).

### 2.2 Eligibility Criteria

The population which was considered for the study was humans of all ages and animal models, and the intervention which was explored was plants and/or their products with anticonvulsant potential. No comparisons were made in this study, and the outcomes for eligibility were studies addressing challenges, barriers, and limitations in translating natural plant anticonvulsants from bench to bedside. There were no limitations on time and setting for studies to be included in the review.

### 2.3 Information Sources and Search Strategy

An initial limited search was conducted in Pubmed using the keywords translational medicine, challenges, plant, anticonvulsant, antiepileptic, and bed-to-bedside. Following the initial search, the text words in the titles and abstracts, and index terms of the retrieved papers were analysed. A second search was then conducted across all included databases—PubMed, Scopus, Google Scholar, and ClinicalTrials.gov—using the identified keywords and index terms. This search was carried out between November 11th and November 20th, 2023. The search in Google Scholar was done via a third-party application called Harzing’s Publish or Perish (Harzing, 2016), where the search results limit was set at 1000 relevant articles - the maximum number of results per search. The reference lists of the sources chosen after full-text review and/or those included in the study were examined to identify additional sources. The complete search strategy can be found in Appendix A.

The articles were imported into Rayyan.ai (Ouzzani et al., 2016) where duplicates were detected and deleted or resolved. The articles were then screened by the authors.

### 2.4 Selection of Sources of Evidence

The authors conducted a pilot test by autonomously screening five publications and subsequently convened to discuss each publication and resolve any uncertainties. A blind screening of the titles and abstracts commenced, followed by a blind full-text screening of eligible articles by the authors in Rayyan.ai to identify potentially relevant articles for this review. All conflicts which arose during the screening process were arbitrated by the lead author for a resolution to be made.

### 2.5 Data Charting Process

A standardised data abstraction tool designed for this study was developed in Microsoft Excel, and this was used to chart data from all eligible studies iteratively by 2 reviewers. Verification and conflict resolutions were made by the lead author. Relevant data that addressed the challenges that hinder the successful translation of natural plant anticonvulsants from preclinical research to clinical application anywhere in the articles were captured during data abstraction charting.

### 2.6 Data Items

Data on characteristics of the studies, such as the author, title of the article, country of origin, year of article publication and institution affiliation of the lead author, as well as challenges that hinder the successful translation of natural plant anticonvulsants from preclinical research to clinical application were abstracted.

### 2.7 Synthesis of Results

A descriptive statistical analysis was performed on data related to the characteristics of the studies. Data on the challenges that hinder the successful translation of natural plant anticonvulsants from preclinical research to clinical application were synthesised through a descriptive qualitative content analysis method (Peters et al., 2020). An inductive approach which was collaborative and iterative was used to synthesise and categorise these data. This was done by the review team at two online meetings to examine each of the challenges from the 37 included articles, to discuss, to code extracted statements and to categorise coded statements based on similarities. Figure 1 provides an example of such statements.

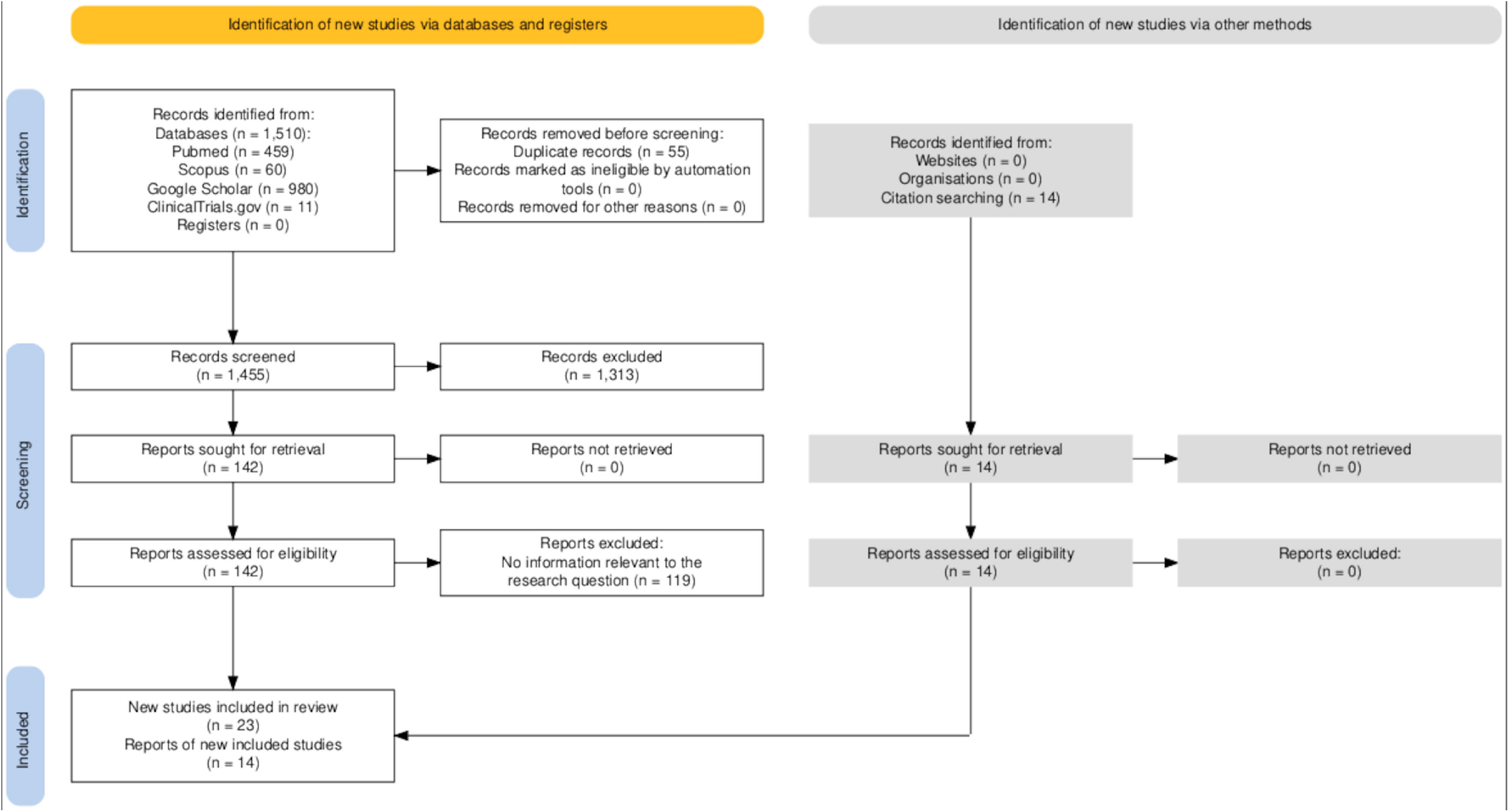

## 3. RESULTS

After searching the four databases, 1,510 articles were obtained from which 55 articles were deleted as duplicates. Based on the title and abstract screening, 1,313 articles were deleted, thus 142 articles proceeded to the full-text screening stage. At this stage, 119 articles were excluded on the basis that they contained no information relevant to the research question. Thus, 23 articles were included at the end of the full-text screening process. The reference list of these 23 articles was critically searched to identify potential articles relevant to the research topic. At the end of the reference screen, an additional 14 relevant articles were identified and retrieved. Thus, a total of 37 articles met the eligibility criteria for this review. Figure 2 (Prisma Flow Diagram) illustrates the details of the selection process and reasons for exclusion, and the extraction datasheet for the included articles can be found in Appendix B.

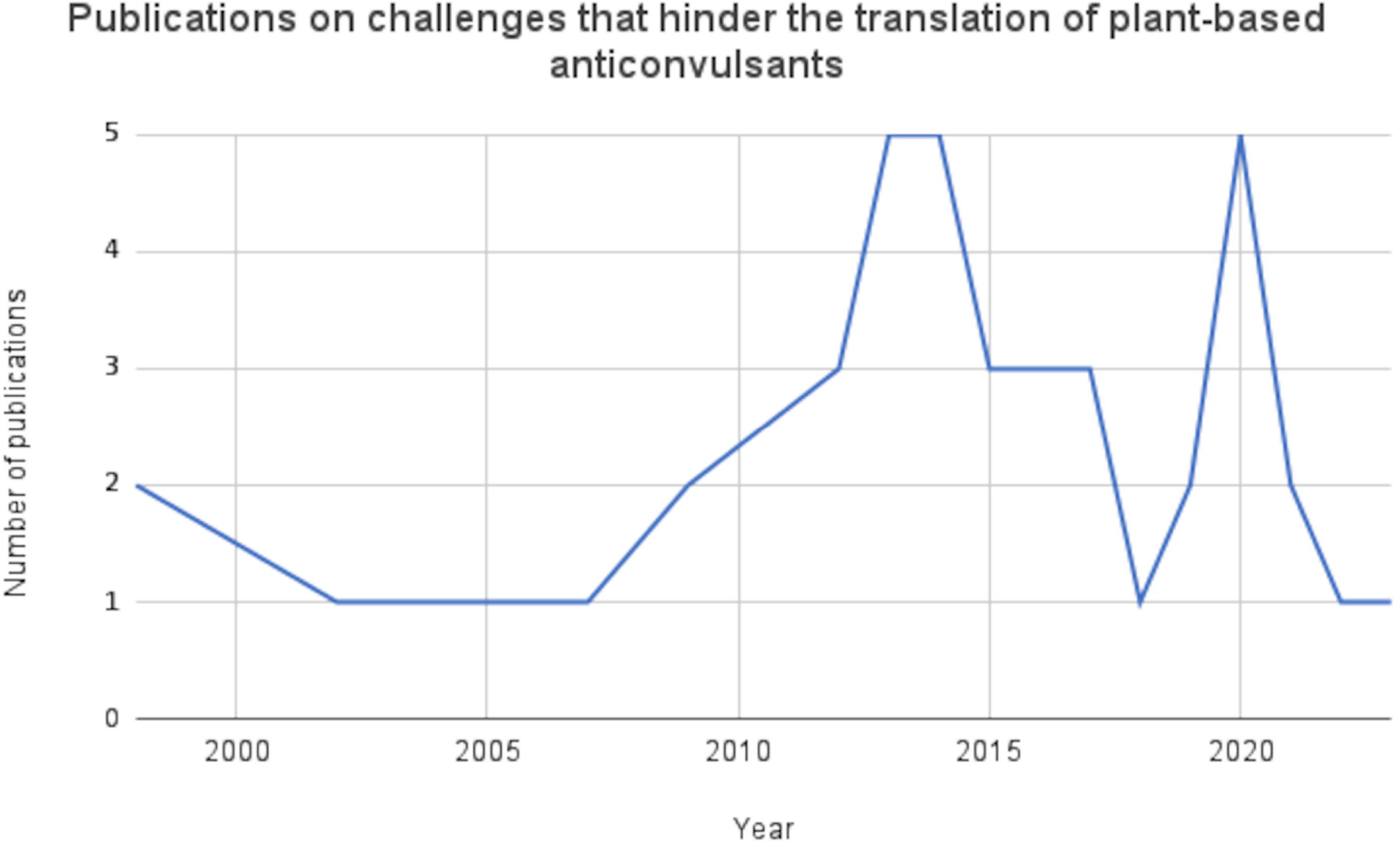

### 3.1 Characteristics of included articles

As illustrated in Figure 3, there are noticeable fluctuations in the number of articles published per year that addressed challenges that hinder the translation of plant-based anticonvulsants from bench to bedside The peak years were 2014 and 2020 with about 14 per cent (n=5) of the included articles being published in each of those years. Almost half of the included studies were carried out in the North American region (n=16; 43.2%) as depicted in Table 1.

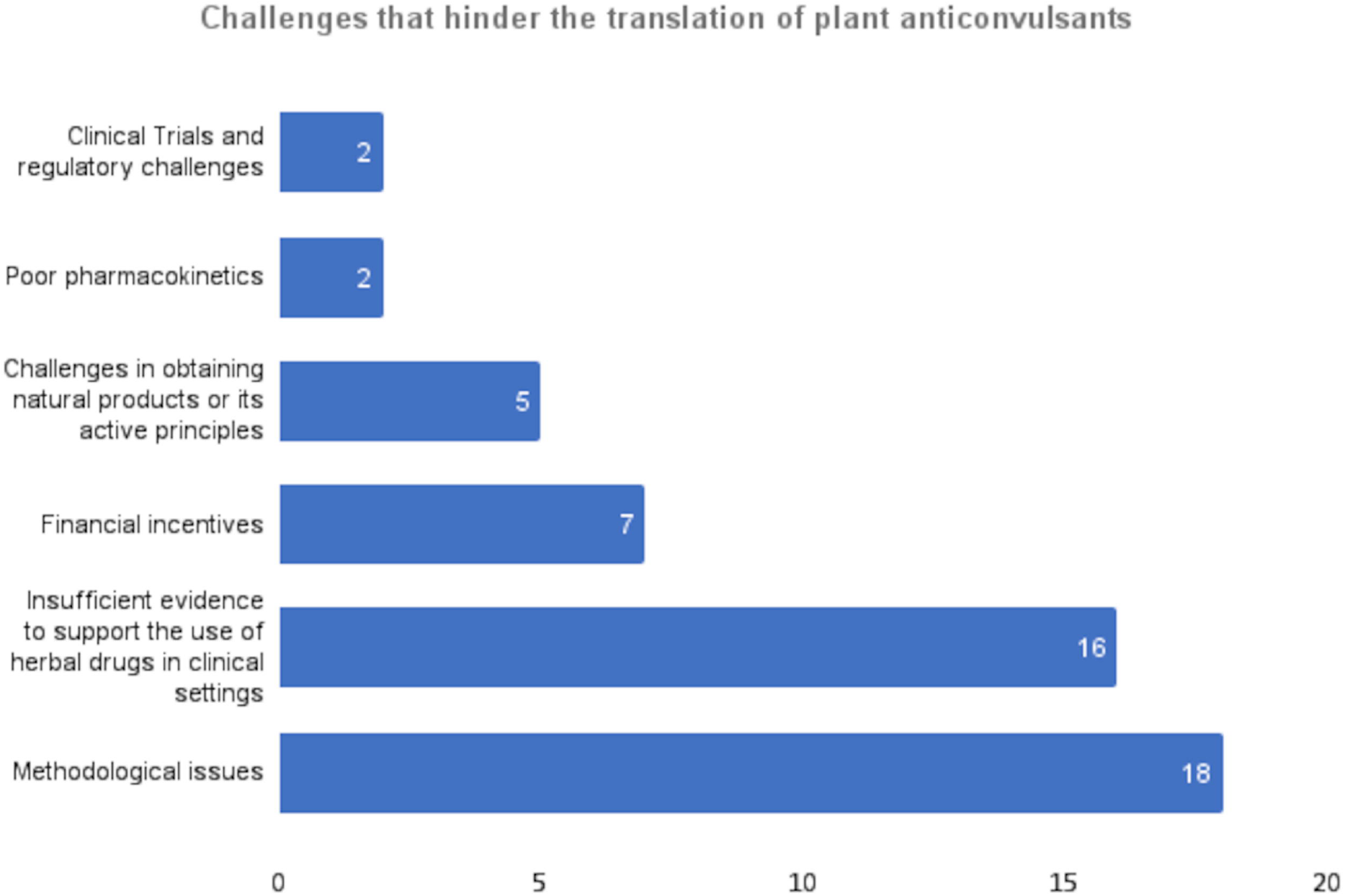

**Table 1.** Regions where studies were conducted.

| **Regions** | **Number of Articles** |
| --- | --- |
| **North America** | 16 |
| **Europe** | 10 |
| **South America** | 4 |
| **Asia** | 4 |
| **Africa** | 1 |
| **Middle East** | 1 |
| **Australia** | 1 |

### 3.2 Challenges that hinder the translation of plant anticonvulsants from bench to bedside

The challenges that hinder the successful translation of natural plant anticonvulsants from preclinical research to clinical applications which were identified in the included studies were analysed inductively. Six distinct categories of challenges were developed with some of the included articles fitting into more than one category. This is presented in Figure 4.

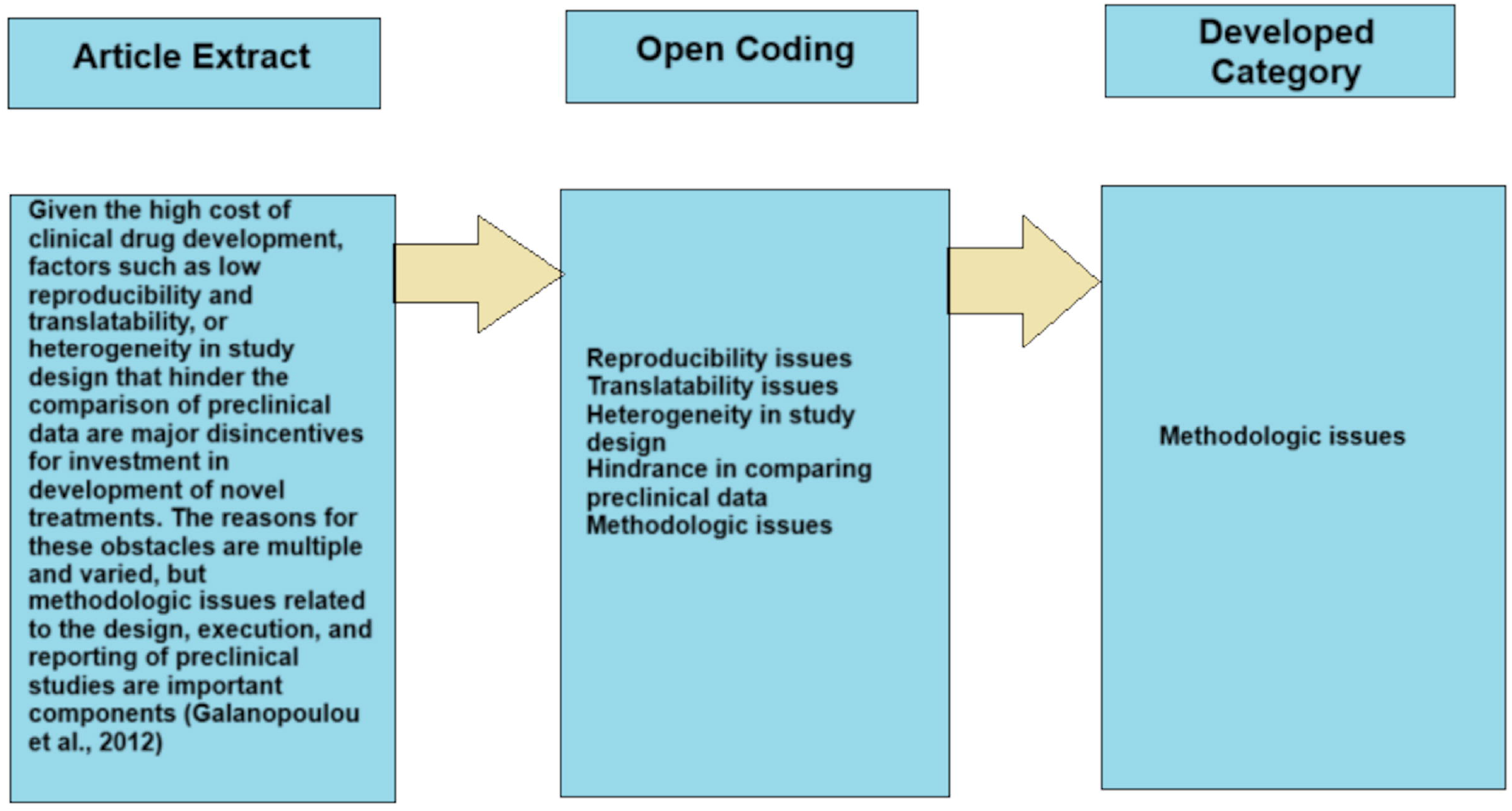

#### 3.2.1 Methodological Issues

The most reported challenge was issues related to methodology in preclinical studies (n=17).

> *Given the high cost of clinical drug development, factors such as low reproducibility and translatability, or heterogeneity in study design that hinder the comparison of preclinical data are major disincentives for investment in the development of novel treatments. The reasons for these obstacles are multiple and varied, but methodologic issues related to the design, execution, and reporting of preclinical studies are important components* (Galanopoulou et al., 2012).

Some of the methodological issues raised by these papers included the failure to incorporate detailed traditional use (e.g. posology, observed effects, and side effects) in the research design, the choice of workflow (e.g. *in vitro* before *in vivo*), the limited use of experimental models with good translational value (Elisabetsky, 2022), lack of standardisation in preclinical experimental designs and data collection (Harte-Hargrove et al., 2017), lack of blinding and randomization in clinical trials, poor control of experimental conditions (e.g. animal body temperature) (Pitkänen et al., 2013), poor methods of assessing experimental outcomes (Chapman et al., 2012), short follow-up periods for clinical trials, unreported sample size calculations and small sample sizes (Li et al., 2009), misemployment of the concentration–effect paradigm, the over-interpretation of data obtained *in vitro* (Gertsch, 2009), inadequately powered studies, publication and other biases, and lack of study design rigour (Simonato et al., 2013). One systematic review (Li et al., 2009) also raised issues about the heterogeneity of study populations and clinical heterogeneity due to differences in herbal formulations which hinder the comparison of studies of herbal medicines for epilepsy.

#### 3.2.2 Insufficient Evidence to Support the Use of Herbal Drugs in Clinical Settings

As identified, the second most prevalent reason why plant anticonvulsants are not successfully translated into the clinical setting is the lack of sufficient evidence to support the use of these anticonvulsants in the clinical setting (n=15).

> *… despite abundant preclinical data on the anticonvulsant properties of many herbal remedies, there are very few human studies assessing safety and efficacy of these products in epilepsy* (Ekstein, 2015).

The current evidence is insufficient to evaluate the safety, efficacy, dosage and pharmacokinetics of herbal anticonvulsants due to the lack of well-designed, controlled, longitudinal and double-blind studies like randomised controlled trials (Szaflarski & Bebin, 2014).

One author stated *“The clinical study of botanicals in people with epilepsy for seizure control is far less developed than the basic science, and the evidence is generally anecdotal and of poor quality”* (Schachter, 2015).

This makes authorization and prescribing of plant anticonvulsants to patients with epilepsy difficult (Huntsman et al., 2020; Zafar et al., 2021).

In addition, there is a dominant pattern of small sets of experiments on a wide range of plant anticonvulsant species (obviously excluding the best-known and/or important commercialised species) (Elisabetsky, 2022). This lack of extensive data for most plant species does not make a strong case for drug development.

> *… the lack of accumulated data, including a battery of in vivo and in vitro tests, phytochemistry, and toxicology, certainly does not help to make a compelling case for drug development* (Elisabetsky, 2022).

#### 3.2.3 Financial Disincentives

Financial disincentives were also identified as a barrier that hinders the translation of plant anticonvulsants into the clinical setting (n=7).

> … *unlike synthetic molecules, whole plants could not be patented, thus denying the pharmaceutical industry a major financial incentive for development* (Gillis, 1998).
>
> … *the unpredictable nature of epilepsy often means lengthy clinical trials. Clinical development for antiepileptic drugs is inevitably more complex and time-consuming than for many other drugs. As a result, the interval between product approval and loss of patent is often shorter* (Schwabe, 2002).

Many in the industry view the epilepsy therapeutics market as saturated. (Simonato M et al., 2014). As a result, there is a significant barrier to demonstrating that a new drug genuinely offers added value, such as improved efficacy. (Sucher & Carles, 2015). The burden of new anticonvulsants to demonstrate superior efficacy to already existing anticonvulsants, in addition to the difficulty in obtaining patents for natural plants, the unpredictable nature of epilepsy which often leads to a more complex, time-consuming, and high-failure-rate clinical development than for many other drugs, deters many in the pharmaceutical industry from investing in the development of plant anticonvulsants. The shorter interval between product approval and patent loss due to the inevitably time-consuming clinical development of plant anticonvulsants also compounds the problem.

#### 3.2.4 Challenges in Obtaining Natural Products or its Active Principles

The challenge of obtaining natural products or its active principles either for use by researchers in drug development studies or patients for use poses a huge challenge to the successful application of plant anticonvulsants. This was recognized by five of the included studies.

> *Some major limiting factors along the way of drug discovery programs from plant sources include the often small quantities of isolated phytochemicals for screening purposes, the labour-intensive and time-consuming protocol of extraction and bioassay-guided fractionation of plant materials, compounded by the fact that forests, especially in Africa, are slowly disappearing and hence plant species are being threatened by extinction* (Ntie-Kang et al., 2014).

One of the studies mentioned Ames and Cridland’s report in a “letter to the editor” where they stated their inability to complete their randomised control trial and cross-over study of cannabidiol in patients with poorly controlled epilepsy because they ran out of supply of cannabidiol (Szaflarski & Bebin, 2014).

Deforestation, especially in Africa, varying concentrations of active principles according to growing conditions, the typically limited amounts of isolated phytochemicals available for screening, and the labour-intensive, time-consuming processes of extraction and bioassay-guided fractionation of plant materials all play a role in this problem (Ntie-Kang et al., 2014; Szaflarski & Bebin, 2014).

#### 3.2.5 Poor Pharmacokinetics

Poor pharmacokinetics of plant products was also identified as a limiting factor in the anticonvulsant drug discovery process from plant sources in two of the included studies. This problem often leads to unacceptable levels of toxicity (Ntie-Kang et al., 2014) or poor bioavailability of plant-derived anticonvulsants (Mishra et al., 2021) which impedes the drug discovery process.

> *… the plant-derived natural products are sometimes faced by unacceptable levels of toxicity due to poor pharmacokinetic properties* (Ntie-Kang et al., 2014).

For example, even though there is enough preclinical evidence to support the neuroprotective role of chrysin, a herbal bioactive molecule, in epilepsy, limited clinical studies exist to support its use due to the poor bioavailability of the compound. The low bioavailability (less than 1%) is mainly attributed to its poor aqueous solubility, as well as its extensive presystemic or first-pass metabolism. The major portion of administered chrysin remains unabsorbed and is excreted in faeces, resulting in its poor bioavailability (Mishra et al., 2021).

#### 3.2.6 Clinical Trials and Regulatory Challenges

The development of plant anticonvulsant drugs is sometimes faced with clinical trial and regulatory challenges which hinder their successful translation into the clinical setting. This was discovered in two of the included studies.

Some of the regulatory challenges faced include a generally lower risk tolerance for new drugs and recent class labelling concerning safety signals (such as suicide) that have impacted opportunities in non-epilepsy indications and affected the overall value proposition for antiepileptic medications. Additionally, new antiepileptic drugs may require long-term safety data commitments across various age groups, with paediatric investigational plans mandating the development and testing of new formulations for very young patients (infants aged ≥1 month). Furthermore, commercialization models suggest that an adjunctive indication alone for a marginally differentiated product is insufficient, and promoting the product for additional uses necessitates establishing those specific indications in the labelling. Moreover, the early incorporation of antiepileptic drugs into the epilepsy treatment paradigm as monotherapy is hindered by the need to obtain prior approval as an adjunctive therapy, resulting in significant delays in the discovery of new antiepileptic medications. (Löscher et al., 2020).

The Oregon Health and Science University terminated a phase II randomised, placebo-controlled, double-blind, cross-over clinical trial to test the safety and potential anticonvulsant efficacy of a botanical extract from *Passiflora incarnata* in patients with partial onset epilepsy because of lack of enrolment of participants in the study (Oregon Health and Science University, 2019).

## 4. DISCUSSION

This scoping review aimed to identify the challenges that hinder the translation of plant anticonvulsants to the clinical setting. After reviewing 1,525 articles and filtering out duplicates and irrelevant studies, 37 articles met the eligibility criteria. From these articles, six distinct categories of challenges were identified: (1) methodologic issues; (2) insufficient evidence to support use of herbal drugs in clinical settings; (3) financial disincentives; (4) challenges in obtaining natural products or its active principles; (5) poor pharmacokinetics; (6) clinical trials and regulatory challenges.

Methodological issues in preclinical studies significantly hinder the progression of plant anticonvulsant compounds from academic laboratories to industrial development programs, and ultimately to clinical trials. These issues usually result in poor reproducibility and translatability of preclinical data in the proceeding stages of drug development, as well as heterogeneity of study designs which makes comparison of preclinical data from different settings very difficult. Several factors which are broadly related to the design, execution, and reporting of preclinical studies (Galanopoulou et al., 2012) contribute to issues with methodology.

Despite considerable advancements in the field of epilepsy to translate preclinical experimental models, practices, and procedures into targeted treatments, challenges continue to exist due to a continuous lack of standardised experimental designs and data collection methods. Inconsistencies in blinding and randomization, experimental conditions, assessment of experimental outcomes, sample size calculation and estimation, follow-up periods for clinical trials, study populations and composition of herbal formulations create challenges in interpreting data and comparing results across different studies, laboratories, and research institutions. The resulting difficulty in synthesising data has often made it challenging to interpret findings, which delays the progression of research from preclinical to clinical stages, increases the duration and cost of clinical trials, and prolongs the development of plant-based epilepsy treatments. (Harte-Hargrove et al., 2017).

The lack of integration of detailed traditional use (such as posology, observed effects, and side effects) into the research design, the selection of workflows (for instance, conducting in vitro studies before in vivo), and the limited use of experimental models with high translational value often result in inefficient allocation of time and resources (Elisabetsky, 2022). Pharmacodynamically, it is anticipated that long-term use may produce effects that differ significantly from those observed with acute or sub-chronic administrations at any given molecular target. However, in the planning, screening, or drug development programs, traditional posology is seldom considered, despite its critical importance for CNS drugs since it has been shown that long-term effects on initial targets are often necessary to achieve a new functional state. Furthermore, plant remedies are usually a mixture of compounds that contribute to the anticonvulsant effects that are observed traditionally. Hence, the choice of *in vitro* studies, which are target-driven screenings, for specific active compounds and/or mechanisms of action, before *in vivo* studies may lead to the loss of truly innovative plant anticonvulsants in initial *in vitro* screenings (Elisabetsky, 2022).

Unfortunately, the prevailing pattern consists of a limited number of experiments conducted across a broad range of species, obviously excluding the most well-known and/or commercially significant species (Elisabetsky, 2022), and the data that exist are usually low-powered studies. Pertaining to this, of prominence is insufficient data collected through randomised control trials. Data is lacking on the efficacy, safety and dosing of plant anticonvulsants to support their use in the clinical setting. All these issues make authorization of plant anticonvulsants, prescribing, and accumulation of data for further drug development difficult, thus hindering their progression to the clinical setting.

Drug development in the field of epilepsy is perceived to be a saturated market by most pharmaceutical companies (Simonato M et al., 2014). This puts a burden on novel drugs to prove superior efficacy to already existing drugs to attract financial investment (Sucher & Carles, 2015). The inability to demonstrate superior efficacy may sometimes lead to the abandonment of a lot of preclinical plant anticonvulsants, subsequently hindering their progression to the clinical setting. Furthermore, the fact that whole plants, unlike synthetic molecules, cannot be patented, in addition to the short interval between product approval and loss of patent due to unusually lengthy clinical trials caused by the unpredictable nature of epilepsy makes investment in plant anticonvulsant drug development unattractive (Gillis, 1998; Schwabe, 2002).

The protocol of extraction and bioassay-guided fractionation of plant materials is usually labour-intensive and time-consuming, and often leads to small quantities of isolated phytochemical compounds for research purposes. Concentrations of plant active principles also vary with different growth conditions such as variable seasons, temperature and weather conditions which sometimes limit the quantities of active principles. Additionally, deforestation, especially in Africa, threatens plant species with extinction. This hugely affects the availability of plant anticonvulsants to patients, and also affects the quantities available for research purposes, to the extent of causing the termination of some clinical trials (Szaflarski & Bebin, 2014).

Poor pharmacokinetic properties of plant anticonvulsants which may result in unacceptable toxicity levels and poor bioavailability sometimes impede the development of plant anticonvulsants. Additionally, the stringent regulatory safety requirements for new anticonvulsants, the need for new anticonvulsants to be first approved as adjunctive treatments before gaining approval as monotherapy medications, and the lack of participants during clinical trials also create challenges in the drug discovery process of plant anticonvulsants.

## 5. CONCLUSION

Epilepsy, a chronic neurological disorder, continues to affect about 50 million people worldwide. Even though efforts have been made to discover alternative anticonvulsants from plants because of their historical use in traditional medicine, most plant anticonvulsants that have been studied have not been able to successfully reach the clinical setting. This study sought to identify the various challenges that affect the translation of plant anticonvulsants from the bench to the bedside. The challenges that were identified include issues related to research methodology, financial disincentives, insufficient evidence to support the use of herbal drugs in clinical settings, challenges in obtaining natural products or their active principles, poor pharmacokinetics, stringent clinical trials and regulatory challenges. Understanding and effectively addressing these challenges will ensure that more plant-based anticonvulsants are successfully translated into clinical practice, thereby enhancing the treatment of epilepsy. Recommendations have been made to tackle some of these challenges that plant anticonvulsants face in the drug development process.

## 6. RECOMMENDATIONS

To overcome issues related to methodology as a barrier to the successful progression of plant anticonvulsants from the bench to bedside, it is imperative to develop and adopt standardised protocols for experimental design, data collection and reporting. This can be facilitated by promoting the usage of common data elements (CDEs). CDEs are standardised terms used to collect and record data as a way to systematically collect information essential to research (Harte-Hargrove et al., 2017). The efforts of the AES/ILAE Translational Research Task Force of the ILAE towards making a publicly available database of CDEs to be used in preclinical epilepsy research are very commendable. This will not only standardise data from epilepsy research but also provide a database of accumulated data, facilitating data analysis and expediting the transition from epilepsy research to the development of treatments for the condition.

Additionally, more adequately powered studies and randomised controlled trials need to be conducted in order to make a compelling case for the use of plant anticonvulsants in the clinical setting (Szaflarski & Bebin, 2014). We also recommend frequent reviews and revisits of the developments in a patenting policy of plant anticonvulsants and their patenting strategy which will help attract inventors and better protect their inventions (Wong & Chan, 2014).

## 7. LIMITATIONS AND STRENGTHS

The limitations of this review are primarily related to the extent of the search. The use of a third-party application (Harzing’s Publish or Perish) for the Google Scholar search might have led to a biased selection of articles. This can impact the comprehensiveness and representativeness of the included studies. Additionally, the keyword terms which were generated for the search in the various databases may have omitted research articles that address the research question which may be described differently with other words. Articles that were published in languages other than English were also excluded. As the quality of the studies was not evaluated, the reliability of the data extracted from the selected studies was not addressed.

The strengths of this review include the meticulousness and transparency in the selection of the included articles. The reporting of this research according to the PRISMA-ScR guidelines (Tricco et al., 2018) also adds to the strength of this study.

## Data Availability

All data produced in the present work are contained in the manuscript

## 8. APPENDICES

Appendix A

Appendix B

## 9. AUTHOR CONTRIBUTIONS

**Joana Opoku:** Conceptualization, Methodology, Formal Analysis, Investigation, Writing - Original Draft, Writing - Review and Editing. **Patrick Amoateng:** Conceptualization, Methodology, Formal Analysis, Investigation, Writing - Original Draft, Writing - Review and Editing, Project Administration. **Kennedy Kwami Edem Kukuia:** Writing - Review and Editing. **Samuel Ankamah:** Writing - Review and Editing. **Emelia Oppong Bekoe:** Writing - Review and Editing. **Samuel Adjei:** Writing - Review and Editing. **Dorcas Osei-Safo:** Writing - Review and Editing. **Samuel Binamin Kombian:** Writing - Review and Editing.

