## Appendix A for "Challenges Hindering the Translation of Plant Anticonvulsants from Bench to Bedside: A Scoping Review"

**APPENDIX 1: SEARCH STRATEGY**

Keywords

| Keyword | Alternative search words |
| --- | --- |
| Bench to bedside | "Translational medicine" OR "Translational research" OR "Translational science" OR "bench to bedside" OR "bench-to-bedside" OR "bedside to bench and back" OR “Drug development” OR “drug discovery” |
| challenges | challenges OR obstacles OR difficulties OR barriers OR hindrances OR problems OR issues OR setbacks |
| plant | plant OR botanical OR herbal OR plant-derived OR plant-based OR medicinal OR plant-origin OR ethnomedicinal |
| anticonvulsant | anticonvulsants OR "Anticonvulsant Agents" OR "Anticonvulsant Drugs" OR antiepileptics OR "Antiepileptic Agents" OR "Antiepileptic Drugs" OR seizure OR epilepsy OR convulsion |

**Search in Pubmed**

| Search Number | Query | Results |
| --- | --- | --- |
| #1 | ((((((("Translational medicine") OR ("Translational research")) OR ("Translational science")) OR ("bench to bedside")) OR ("bench-to-bedside")) OR ("bedside to bench and back")) OR ("Drug development")) OR ("drug discovery") | 341,887 |
| #2 | (((((((challenges) OR (obstacles)) OR (difficulties)) OR (barriers)) OR (hindrances)) OR (problems)) OR (issues)) OR (setbacks) | 3,787,290 |
| #3 | (((((((plant) OR (botanical)) OR (herbal)) OR (plant-derived)) OR (plant-based)) OR (medicinal)) OR (plant-origin)) OR (ethnomedicinal) | 9,683,868 |
| #4 | ((((((((anticonvulsants) OR ("Anticonvulsant Agents")) OR ("Anticonvulsant Drugs")) OR (antiepileptics)) OR ("Antiepileptic Agents")) OR ("Antiepileptic Drugs")) OR (seizure)) OR (epilepsy)) OR (convulsion) | 432,223 |
| #5 (#1 AND #2 AND #3 AND #4) | (((((((((("Translational medicine") OR ("Translational research")) OR ("Translational science")) OR ("bench to bedside")) OR ("bench-to-bedside")) OR ("bedside to bench and back")) OR ("Drug development")) OR ("drug discovery")) AND ((((((((challenges) OR (obstacles)) OR (difficulties)) OR (barriers)) OR (hindrances)) OR (problems)) OR (issues)) OR (setbacks))) AND ((((((((plant) OR (botanical)) OR (herbal)) OR (plant-derived)) OR (plant-based)) OR (medicinal)) OR (plant-origin)) OR (ethnomedicinal))) AND (((((((((anticonvulsants) OR ("Anticonvulsant Agents")) OR ("Anticonvulsant Drugs")) OR (antiepileptics)) OR ("Antiepileptic Agents")) OR ("Antiepileptic Drugs")) OR (seizure)) OR (epilepsy)) OR (convulsion)) | 459 |

**Search in Scopus**

Title/abstract/keyword search:

("Translational medicine" OR "Translational research" OR "Translational science" OR "bench to bedside" OR "bench-to-bedside" OR "bedside to bench and back" OR “Drug development” OR “drug discovery”)

AND

(challenges OR obstacles OR difficulties OR barriers OR hindrances OR problems OR issues OR setbacks)

AND

(plant OR botanical OR herbal OR plant-derived OR plant-based OR medicinal OR plant-origin OR ethnomedicinal)

AND

(anticonvulsants OR "Anticonvulsant Agents" OR "Anticonvulsant Drugs" OR antiepileptics OR "Antiepileptic Agents" OR "Antiepileptic Drugs" OR seizure OR epilepsy OR convulsion)

**Search in Google Scholar**

All words anywhere in document:

anticonvulsant plant translation challenges

**Search in ClinicalTrials.gov**

condition/disease: seizure OR epilepsy OR convulsion

Intervention/treatment: (anticonvulsants OR "Anticonvulsant Agents" OR "Anticonvulsant Drugs" OR antiepileptics OR "Antiepileptic Agents" OR "Antiepileptic Drugs" OR seizure OR epilepsy OR convulsion) AND (plant OR botanical OR herbal OR plant-derived OR plant-based OR medicinal OR plant-origin OR ethnomedicinal)
